# Just 2% of SARS-CoV-2-positive individuals carry 90% of the virus circulating in communities

**DOI:** 10.1101/2021.03.01.21252250

**Authors:** Qing Yang, Tassa K. Saldi, Erika Lasda, Carolyn J. Decker, Camille L. Paige, Denise Muhlrad, Patrick K. Gonzales, Morgan R. Fink, Kimngan L. Tat, Cole R. Hager, Jack C. Davis, Christopher D. Ozeroff, Nicholas R. Meyerson, Stephen K. Clark, Will T. Fattor, Alison R. Gilchrist, Arturo Barbachano-Guerrero, Emma R. Worden-Sapper, Sharon S. Wu, Gloria R. Brisson, Matthew B. McQueen, Robin D. Dowell, Leslie Leinwand, Roy Parker, Sara L. Sawyer

## Abstract

We analyze data from the Fall 2020 pandemic response efforts at the University of Colorado Boulder (USA), where more than 72,500 saliva samples were tested for SARS-CoV-2 using quantitative RT-PCR. All samples were collected from individuals who reported no symptoms associated with COVID-19 on the day of collection. From these, 1,405 positive cases were identified. The distribution of viral loads within these asymptomatic individuals was indistinguishable from what has been previously reported in symptomatic individuals. Regardless of symptomatic status, approximately 50% of individuals who test positive for SARS-CoV-2 seem to be in non-infectious phases of the disease, based on having low viral loads in a range from which live virus has rarely been isolated. We find that, at any given time, just 2% of individuals carry 90% of the virions circulating within communities, serving as viral “super-carriers” and possibly also super-spreaders.

## Introduction

SARS-CoV-2 is a novel coronavirus that emerged into the human population in late 2019 [1], presumably from animal reservoirs [2,3]. During the ensuing world-wide pandemic, already more than two million lives have been lost due to the virus. Spread of SARS-CoV-2 has thus far been extremely difficult to contain. One key reason for this is that both pre-symptomatic and asymptomatic infected individuals can unwittingly transmit the virus to others [4–13]. Further, it is becoming clear that certain individuals play a key role in seeding super-spreading events [14– 17]. Here, we analyzed data from a large university surveillance program. Viral loads were measured in saliva, which has proven to be an accessible and reliable biospecimen in which to identify carriers of this respiratory pathogen, and the most likely medium for SARS-CoV-2 transmission [18–20]. Our dataset is unique in that all SARS-CoV-2-positive individuals reported no symptoms at the time of saliva collection, and therefore were infected but asymptomatic or pre-symptomatic. We find that the distribution of SARS-CoV-2 viral loads on our campus is indistinguishable from what has previously been observed in symptomatic individuals. Strikingly, these datasets demonstrate widespread differences in viral levels between individuals, with a very small minority of the infected individuals harboring the majority of the infectious virions.

## Results

### The University of Colorado Boulder SARS-CoV-2 screening operation

We analyzed data resulting from SARS-CoV-2 testing performed on the University of Colorado Boulder (USA) campus during the Fall academic semester of 2020 (Aug. 27^th^ – Dec. 11^th^, 2020). Residents of dormitories were tested weekly, and several campus testing sites were in operation throughout the semester offering testing for any campus affiliate. At the time of saliva collection participants were asked to confirm that symptoms were not present; therefore, any infected persons identified through this surveillance testing were asymptomatic or pre-symptomatic at the time of saliva collection. It should be noted that all of the samples analyzed herein were collected before the B.1.1.7 (“U.K.”) SARS-CoV-2 variant, and subsequent major variants of concern, were first documented in the United States during the final weeks of 2020 and the beginning of 2021 [21].

During the Fall 2020 semester, more than 72,500 saliva samples were screened for SARS-CoV-2. A quantitative reverse transcription-polymerase chain reaction (RT-qPCR) assay was used, with the template coming from the direct addition of saliva without RNA purification [22]. Three TaqMan primer/probe sets were used in a multiplex reaction directed against two regions of the SARS-CoV-2 genome (CU-E and CU-N, where CU stands for the University of Colorado) and a host transcript (CU-RNaseP). For each of the primer sets that detect the viral genome, a standard curve was generated to relate Ct value (cycle threshold) to viral load (virions per mL) in the original saliva sample (**Supplemental Figure S1)**.

From over 72,500 saliva samples screened, 1,405 SARS-CoV-2-positive samples were identified. The vast majority of these positive samples were from unique individuals, because individuals with positive tests were directed into the healthcare system for further testing and care. The distribution of the Ct values of these 1,405 individuals, with each of the two primer sets used, is shown in **Figure 1A**. Overall, the distribution of SARS-CoV-2 viral load fits under a log-normal distribution centered around the geometric mean of 7.3×10^5^ virions/mL (CU-E primers) or 1.5×10^6^ virions/mL (CU-N primers). The highest observed viral load was over 6 trillion (6.1 × 10^12^) virions/mL, which was only observed in 1 individual, while the lowest viral load detected was 8 virions/mL. Thus, surveillance testing demonstrates an extremely wide variation in the viral load in infected but seemingly healthy (asymptomatic) individuals.

**Figure 1.**
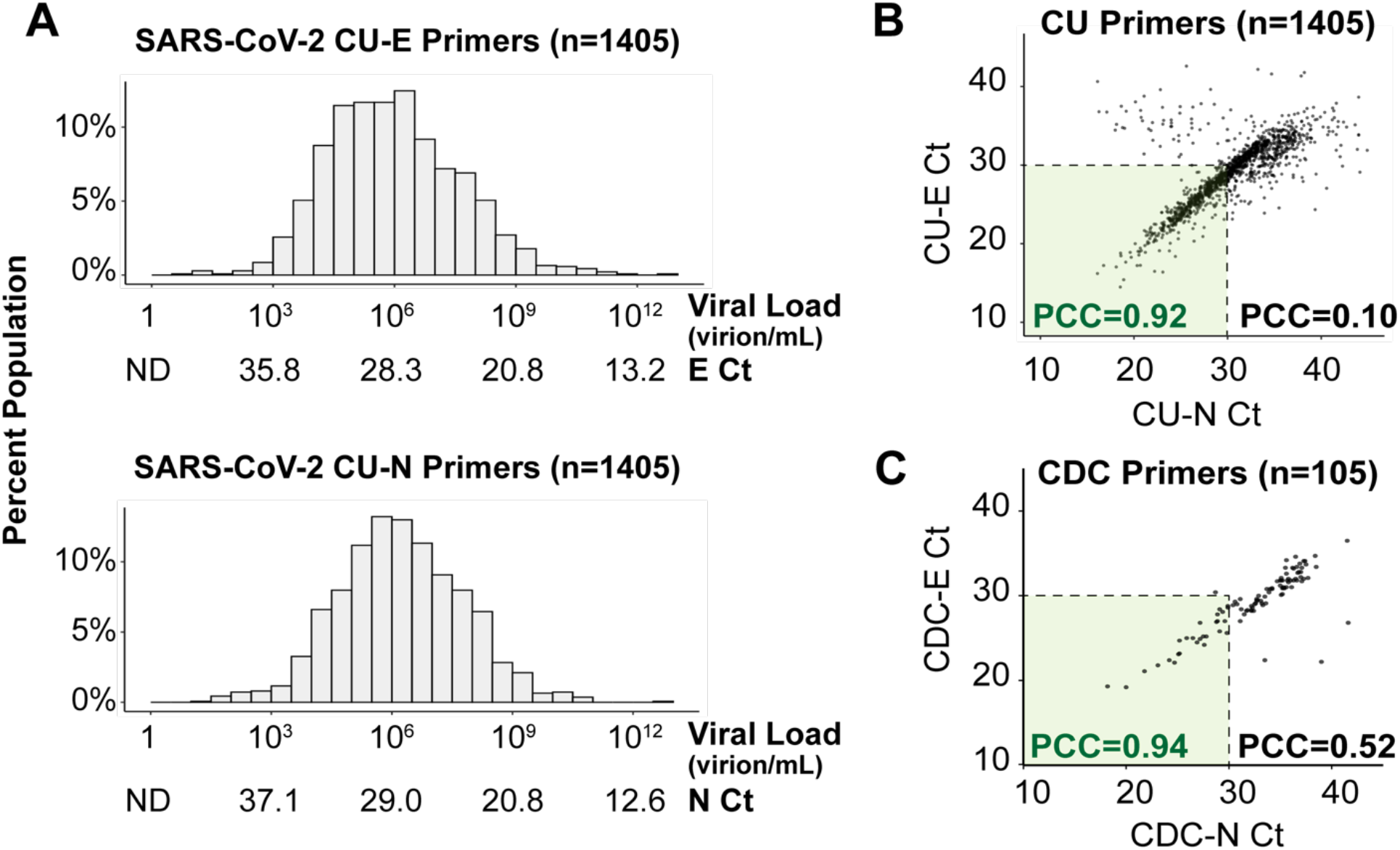
Saliva viral load distribution within our campus population. **A)** Viral load distribution within the 1,405 positive samples identified on campus during the Fall semester of 2020. Each histogram shows Ct values obtained using TaqMan primer/probe sets targeting either the E gene (“CU-E”) or the N gene (“CU-N”) of SARS-CoV-2. The horizontal axes are labeled with both the cycle threshold values (Ct) and the corresponding viral loads calculated from the standard curve for each primer set (**Supplemental Figure S1**). ND = no data, as the viral load is below the RT-qPCR detection limit. **B)** The Ct values resulting from the two primer sets in panel A are highly correlated, especially in samples with high viral loads (Ct value lower than 30). Pearson correlation coefficients (PCC) are shown within and beyond the Ct=30 arbitrary cutoff. **C)** For 105 of the SARS-CoV-2 positive saliva samples, we ran RT-qPCR side-by-side with 8 different primer sets commonly used in SARS-CoV-2 diagnostic tests (**Supplemental Figure S2**). Here, we show the same analysis as in panel B, except with the U.S. Centers for Disease Control’s (CDC) primers targeting the E and N genes (see methods).

To verify that these viral load distributions were not influenced by the specific RT-PCR primers used, we determined the agreement between the CU-N and CU-E primers with regard to the Ct values produced from samples. We find a tight correlation at Ct values < 30 (Pearson correlation coefficient of CU-N and CU-E Ct values = 0.92), but this correlation breaks down at higher Ct values (Pearson correlation coefficient of CU-N and CU-E Ct values = 0.10, **Figure 1B**). At high Ct values (i.e. low viral loads) weaker correlation is likely a result of stochasticity in reverse transcription or initial rounds of PCR. This is supported by an in-depth analysis performed on 105 of the SARS-CoV-2 positive samples, where each sample was analyzed with 8 different primer sets commonly used in SARS-CoV-2 diagnostic tests (**Figure 1C** and **Supplementary Figure S2**). In all instances, we see tight congruence between Ct values generated with different primers at Ct values < 30, and less congruence at higher Ct values. Overall, since the CU-E primer set demonstrated higher agreement with other primer sets during this in-depth comparison (**Supplementary Figure S2)**, we used the Ct values resulting from this primer set to calculate saliva viral loads from this point forward.

### Populations have similar viral load distributions regardless of symptomatic status

We next compared viral loads from individuals on our campus, who had no symptoms at the time of sample collection, to similar viral load measurements taken in saliva of symptomatic individuals. We examined published SARS-CoV-2 RT-qPCR datasets derived from studies of hospitalized (and therefore symptomatic) individuals. We specifically sought studies that assayed saliva and where viral loads were reported, since Ct values are lab- and assay-specific [23]. We identified 404 datapoints that met such criteria, which we collated from the 10 studies listed in **Supplementary Table S1**. Similar to the geometric mean that we observed in our campus asymptomatic population (7.3×10^5^ virions/mL), the viral load in symptomatic patient saliva samples shows a log-normal distribution with a geometric mean of 4.0×10^5^ virions/mL and varied from very high viral loads (9.5×10^10^ virions/mL) to viral loads near the limit of detection (1.3 virions/mL) (**Figure 2A**). We next plotted the cumulative distribution of viral load in both populations (**Figure 2B**). This comparison really represents two extremes: one group is mostly hospitalized, while the other group represents a mostly young and healthy (but infected) college population. Yet, the distributions are extremely similar (two-sided two-sample Kolmogorov-Smirnov test, D statistic = 0.04, p-value = 0.718; (**Figure 2B**)). Therefore, individuals have similar distributions of saliva viral load regardless of symptomatic status, as has also been observed in studies of viral load in anterior nasal or nasopharyngeal swabs [24–29].

**Figure 2.**
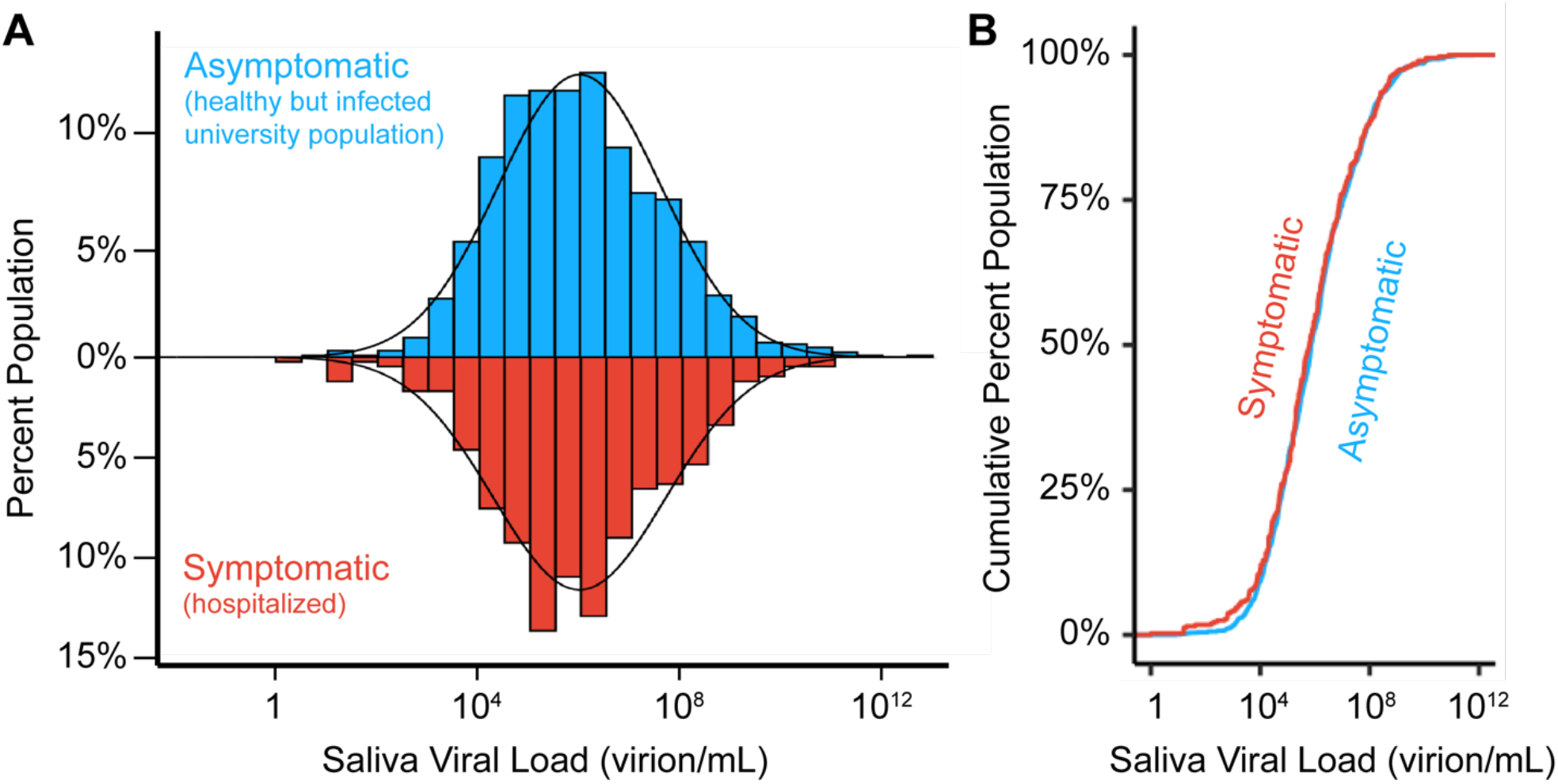
Viral load distributions are similar in asymptomatic and symptomatic populations. **A)** histogram of saliva viral loads in our asymptomatic campus population (N=1405, blue) compared to the same histogram of saliva viral loads from symptomatic (N=404, red) individuals. The latter represents data compiled from the ten studies in **Table S1**. A log-normal probability density function is fitted onto the two distributions given the population mean and standard deviation. **B)** Empirical cumulative distribution functions (ECDFs) of saliva viral load in pre-symptomatic (N=1405, blue) and symptomatic (N=404, red) populations. The similarity of the two ECDFs were assessed with the Kolmogorov-Smirnov test, which resulted in D statistic = 0.04, and p-value = 0.72.

### A small subset of individuals carries most of the circulating virions

We next analyzed how virus is distributed between individuals within populations. By summing the viral load across individuals, starting with those with the highest viral loads, we find that just 2% of individuals harbor 90% of the circulating virions (**Figure 3**). This is true in both the campus (i.e. asymptomatic) and hospitalized (i.e. symptomatic) populations. Further, 99% of community-circulating virions are accounted for by just 12% of the asymptomatic and 14% of the symptomatic population. In both asymptomatic and symptomatic populations, one single individual with the highest saliva viral load carried 5% of the total circulating virions. On the other hand, all individuals with saliva viral loads lower than 10^6^ virions per mL combined (representing ∼50% of the infected individuals) harbor less than 0.02% of the virions in both populations. This can be understood because Ct is a linear representation of logarithmic increases in vial load, so that the viral load increases exponentially as the Ct value decreases (**Supplemental Figure S1**). Thus, there is a highly asymmetric distribution of viruses within both populations, with just a small number of people carrying the vast majority of the virus. It remains unknown whether these are special individuals capable of harboring extraordinarily high viral loads, or whether many infected individuals pass through a very short time period of extremely high viral load (see further discussion below). Irrespective of mechanism it is nevertheless true that, at any given moment in time, a small number of people are harboring the vast majority of virions.

**Figure 3:**
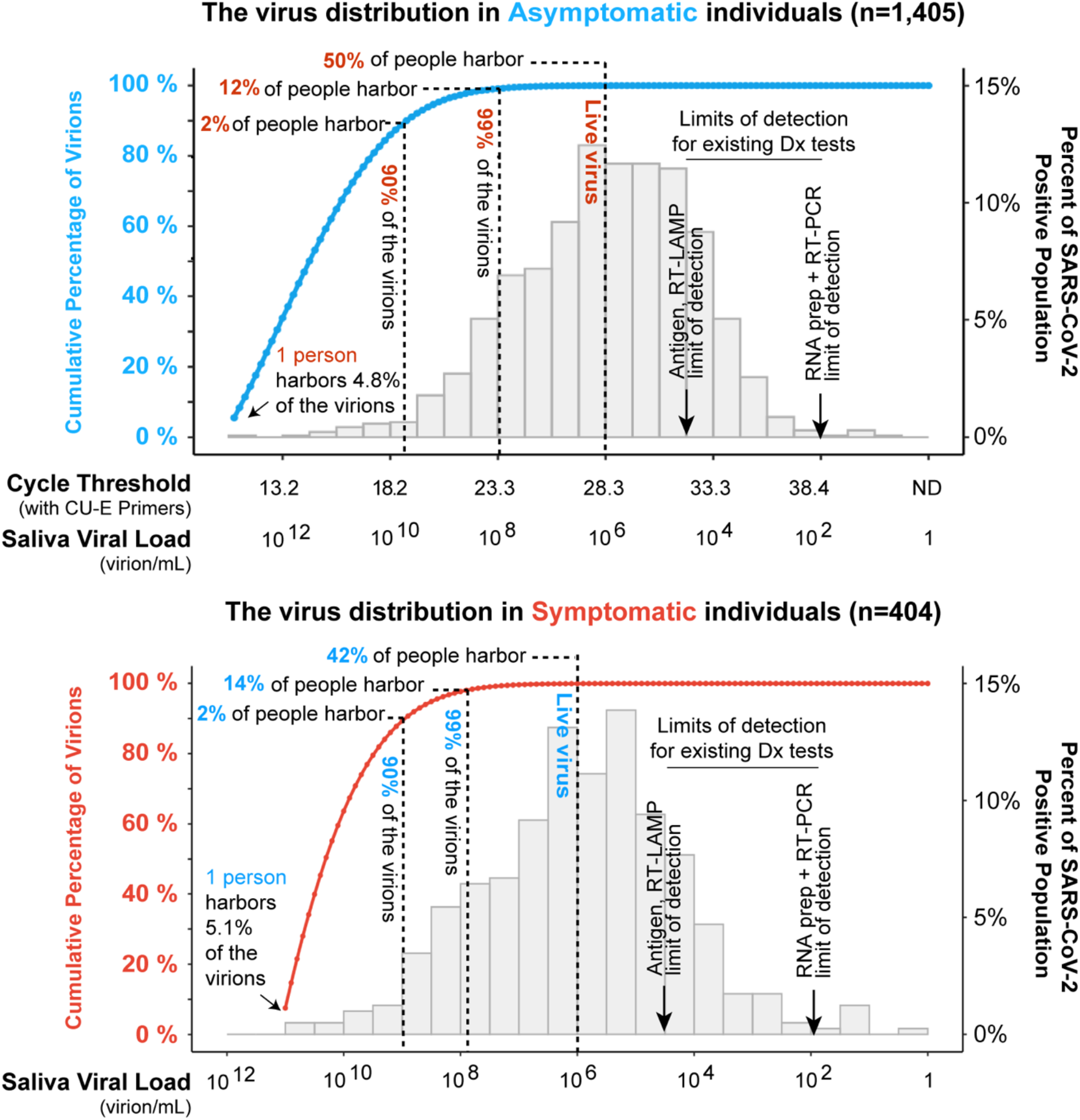
A small percentage of individuals are viral super-carriers. The histograms shown (right Y axes) are the same as were shown in figure 2. Starting from the left of each histogram (i.e. those individuals with the highest viral loads), we calculated the accumulative percentage of total virions as a function of saliva viral load based on the probability density function of the distribution (blue and red lines, and left Y axes). In both asymptomatic (blue line) and symptomatic populations (red line), the portion of population that harbors 90% and 99% of the circulating virus is highlighted by the dashed lines. We estimate that only about 50% of individuals who test positive for the virus actually harbor live virus, based on the observation that live virus has rarely been isolated from samples with viral loads <10^6^ virions per mL [28,30–35]. For context, typical limits of detection of three major SARS-CoV-2 testing paradigms are shown. RT-qPCR on purified RNA is the most sensitive, with RT-LAMP and antigen tests performed directly on biological fluids being less sensitive. However, all testing paradigms will capture virtually all infectious individuals and virions, in pre-symptomatic and symptomatic populations alike. Limits of detection are taken from [36,37].

Infectious virions have rarely been isolated from clinical samples of individuals with viral load less than 10^6^ virions per mL [28,30–35]. One hypothesis is that people in this low range of viral load may simply be shedding viral genomes from damaged tissue that is undergoing repair, and for this reason they may not pose a substantial risk of infecting others. Our distributions suggest that approximately half of the people who test positive may not be infectious to others (**Figure 3**) based on this line of reasoning.

## Discussion

An important finding herein is that the vast majority of circulating virions in communities are found within the bodies of a small number of individuals. These findings corroborate similar trends observed elsewhere [14–17,25]. Although it remains to be seen exactly how transmission probability relates to viral load, a strong implication is that these individuals who are viral super-carriers may also be super-spreaders. Higher viral loads have been shown to increase the probability of transmission to others in China [38], in Spain [39], and between pairs of roommates on our University campus (Bjorkman et al, in prep). A higher rate of spread by viral super-carriers would be consistent with recent contact tracing analyses suggesting that 80-90% of infections are spread by 10-20% of infected individuals [14–17]. A higher rate of spread by viral super-carriers would also be consistent with the surprisingly low transmission rates being reported between roommates (Bjorkman et al, in prep), schoolmates [40,41], and household members [42], which could be explained if only a small fraction of infected individuals have high enough viral loads to facilitate active transmission.

One potential explanation for the differences in viral loads between individuals is that individuals were simply tested at different stages of otherwise similar viral infections. However, longitudinal analyses of individual infections show that peak viral loads vary dramatically between individuals [43–45]. Thus, the parsimonious explanation is that individuals produce different levels of virus. Whether this is due to variation in the immune response, variation in host factors supporting virus replication like ACE2, the specific viral variant infecting, or initial infection site or dose remains to be determined [46–49].

The concentration of a majority of the virus in a small fraction of the population at a given time is a critical observation with actionable conclusions. Community screening to identify viral super-carriers within pre-symptomatic and asymptomatic stages of disease will be important, since these individuals will continue to sustain and drive the epidemic if not located. Finding viral super-carriers will have a disproportionately large impact on curbing new COVID-19 infections, yet individuals without symptoms don’t tend to seek out testing so screening will need to target healthy populations. Modeling approaches show that one of the most important factors in screening for SARS-CoV-2 will be the speed with which infected people receive their test results (also referred to as turnaround time) [50]. The longer it takes for people to receive their results, the more time goes by where they might unwittingly infect others. Therefore, it is imperative that we find virus super-carriers, and inform them of their infection status in a way that is fast, easy, and accessible. Although sensitivities vary between current monitoring and diagnostic paradigms, all are more than capable of finding the majority of infected individuals and the vast majority of infectious virions (**Figure 3**).

## Methods

### Collection of University samples

For sample collection conducted at our university, individuals were asked to fill out a questionnaire to confirm that they did not present any symptoms related to COVID-19, and to collect no less than 0.5 mL of saliva into a 5 mL screw-top collection tube. Saliva samples were heated at 95°C for 30 minutes on-site to inactivate the viral particles for safe handling, and then placed on ice or at 4°C before being transported to testing laboratory for quantitative RT-PCR analysis on the same day.

### Saliva quantitative RT-PCR used for screening saliva samples on the University of Colorado Boulder campus

For quantitative RT-PCR analysis, the university testing team transferred 75 μL of saliva into a 96-well plate that had been pre-loaded with 75 μL 2X TBE buffer supplemented with 1% Tween-20. 5 μL of this diluted sample was then added to a separate 96-well plate containing 15 μL reaction mix composed of: TaqPath 1-step Multiplex Master Mix (Thermo Fisher A28523), nuclease-free water, and triplex primer mix consisting of CU-E, CU-N, and CU-RNaseP primer and probe sets (sequence and concentration specified in the table below; conditions changed slightly during the semester). The reactions were mixed, spun down, and loaded onto a Bio-Rad CFX96 or CFX384 qPCR machine. Quantitative RT-PCR was run using the standard mode, consisting of a hold stage (25°C for 2 minutes, 50°C for 15 minutes, and 95°C for 2 minutes) followed by 44 cycles of a PCR stage (95°C for 3 seconds, 55°C for 30 seconds, with a 1.6°C/sec ramp up and ramp down rate). Ct values from all campus testing efforts were communicated to us as de-identified data.

### Focused analysis of 105 SARS-CoV-2 positive samples

For smaller subsets of samples, as described herein, we did a side-by-side comparison of 3 different quantitative RT-PCR multiplex assays commonly used in SARS-CoV-2 diagnostics. We thawed 105 frozen, de-identified saliva samples which had previously tested positive for SARS-CoV-2 in the campus screening operation. We performed quantitative RT-PCR analysis using different primer probe combinations. First, 25 μL of thawed, previously heat-treated saliva was transferred into a 96-well plate where each well had been pre-loaded with 25 μL 2xTBE buffer supplemented with 1% Tween-20. Next, 5 μL of the diluted sample was added to separate 96-well plates containing 15 μL reaction mix composed of: TaqPath 1-step Multiplex Master Mix (Thermo Fisher A28523), nuclease-free water and CDC triplex primer mix or CU triplex primer mix (sequence and concentration specified in the table below). The reactions were mixed, spun down, and loaded onto a Bio-Rad CFX96 qPCR machine. Quantitative RT-PCR was run using the standard mode, consisting of a hold stage (25°C for 2 minutes, 50°C for 15 minutes, and 95°C for 2 minutes) followed by 44 cycles of a PCR stage (95°C for 3 seconds, 55°C for 30 seconds, with a 1.6°C/sec ramp up and ramp down rate). Each plate also contained wells with 5 μL negative control template containing nuclease-free water diluted 1:1 with 1% Tween-20 and 5 μL positive control template containing synthetic SARS-CoV-2 RNA (Twist Biosciences 102024) diluted to 1000 genome copies/μL and total human reference RNA (Agilent 750500) diluted to 10ng/μL in nuclease-free water.

We also performed the SalivaDirect TaqMan RT-qPCR analysis [20] on each of these samples. 75 μL of each saliva specimen were combined with 9.4 μL of Proteinase K (20 mg/mL, NEB, P8107S). Samples were incubated at ambient temperature for 15 minutes and then heated to 95^0^C for 5 minutes to inactive the Proteinase K. Next, 5 μL of saliva was used as template in a 20 μL reaction that also contained 1X TaqPath 1-step Multiplex Master Mix, nuclease free water and primer and probe sets at concentrations described here. The RT-qPCR was run on the BioRad CFX96 qPCR machine using the same program described for the CU assays. Along with residual saliva samples, a duplicate serial dilution of contrived positive samples made from healthy donor saliva and heat inactivated viral particles were carried through the SalivaDirect protocol [20]. Cycle threshold values from these samples were used to create a standard curve in order to estimate viral load for each residual sample.

### RT-qPCR Taqman primer/probe sets used for university screening and focused analysis

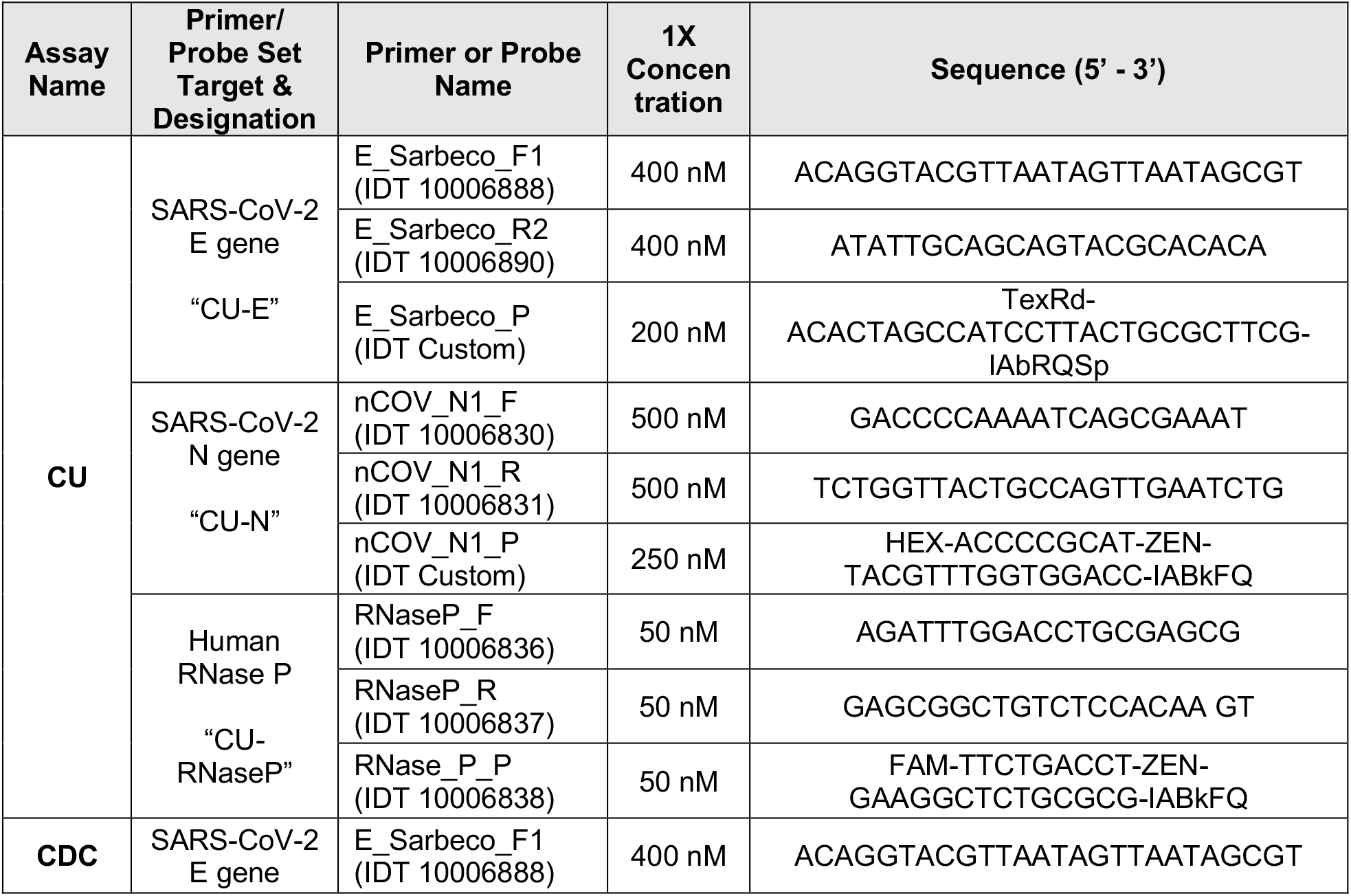
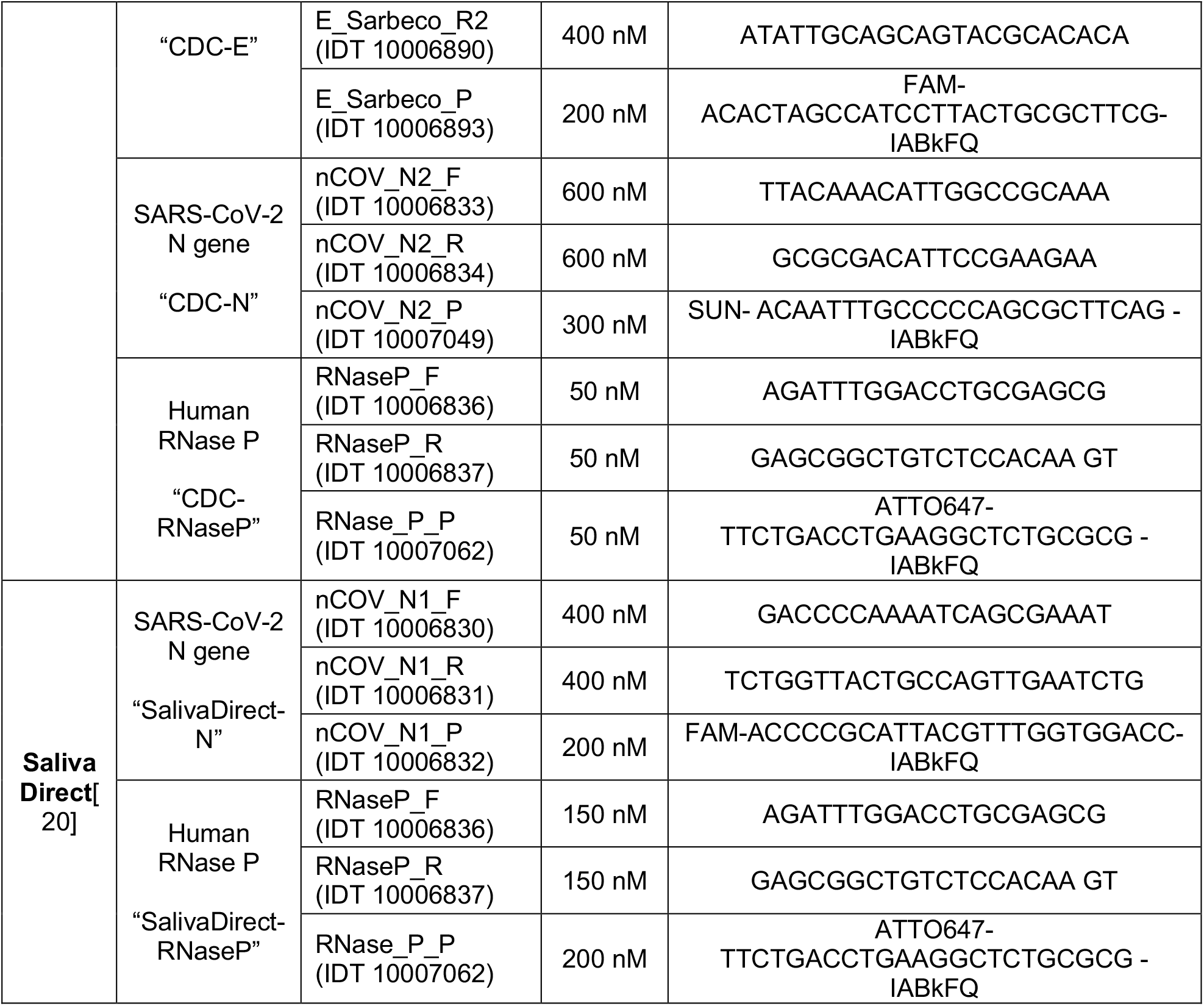

## Data Availability

All data is contained within the manuscript.

## SUPPLEMENTAL INFORMATION

**Supplementary Table S1.**
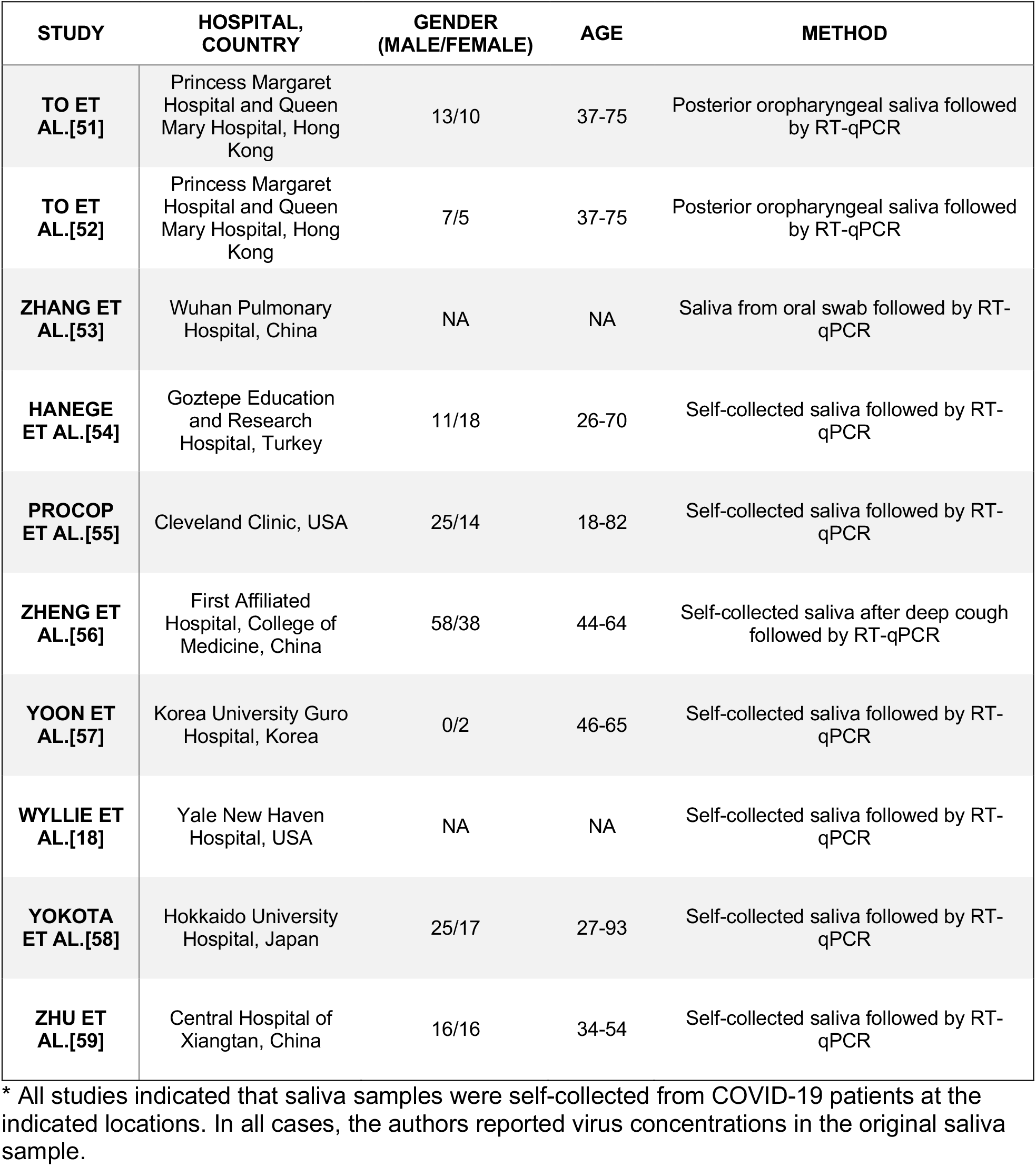
Studies from which viral loads in symptomatic individuals were derived*.

**Figure S1.**
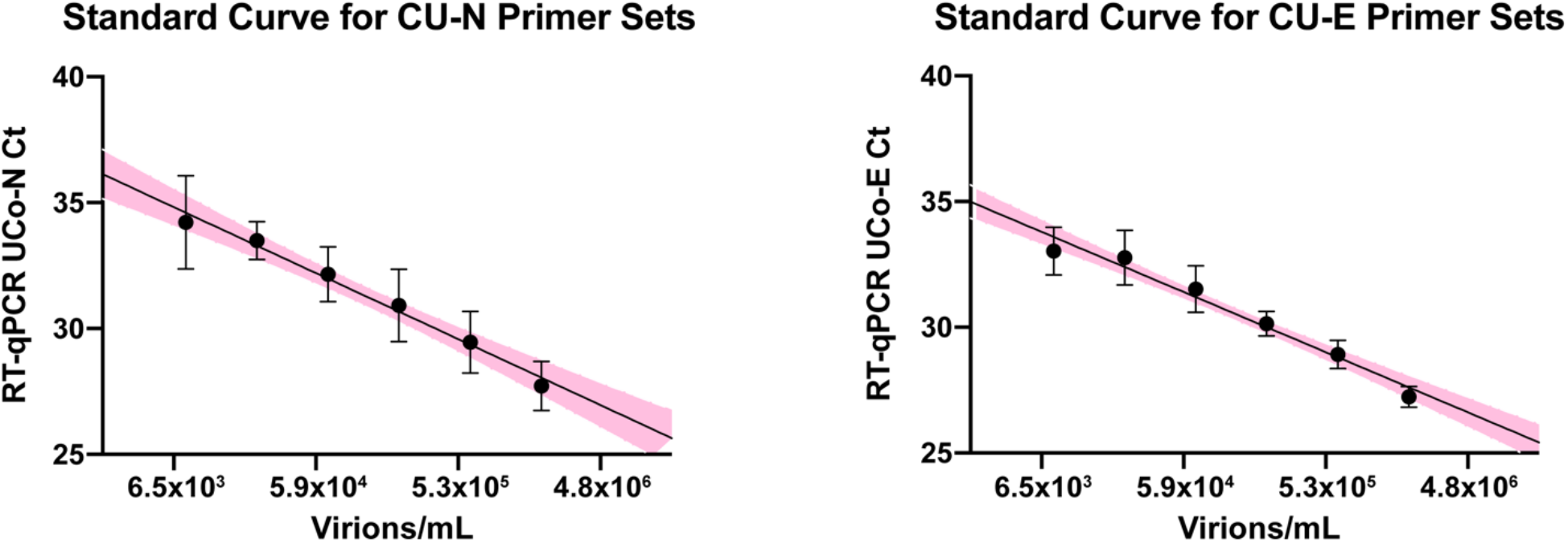
Standard curves for primer sets used in this study. 10,000 virions/µL of heat deactivated SARS-CoV-2 virus was spiked into negative saliva specimens from 6 different individuals and incubated for 30 minutes at 95°C. Samples were diluted to indicated concentrations using heat-treated saliva without SARS-CoV-2 addition from the same individuals. The standard curve generated from the linear regression analysis is illustrated with 95% confidence interval.

**Figure S2.**
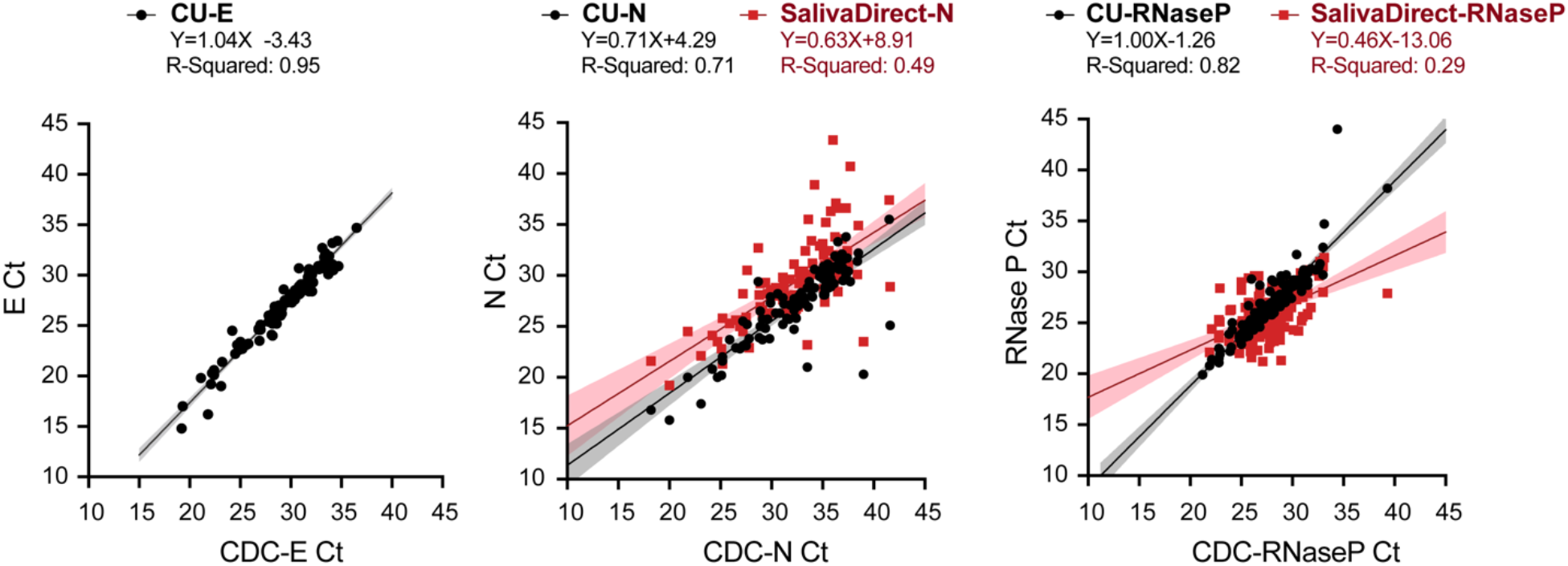
Correlation of Ct values between different primer sets used to quantify saliva viral load. Using 105 SARS-CoV-2 positive saliva samples, we examined the Ct values obtained with different RT-qPCR multiplex assays and compared them via correlation analysis. For 105 virus-positive saliva samples, 8 different Ct values were generated all in one day from each sample, in a side-by-side direct analysis of the performance of each primer set. Ct values from the Centers for Disease Control primers (CDC-E, CDC-N or CDC-RNaseP) are reported on the X-axes. On the Y-axes are plotted the corresponding Ct values resulted from our university screening primers (CU-E, CU-N or CU-RNaseP) and primer sets used in the SalivaDirect [20] test (SalivaDirect-N and SalivaDirect-RNase P) primer sets.

## Ethics statements

Viral load data on student participants were deidentified and aggregated from the University of Colorado Boulder operational screening for SARS-CoV-2. This activity does not meet the definition of human subject research described in the United States Health and Human Services 45 Code of Federal Regulations Part 46. Deidentified saliva samples (n=105) used for the cross-comparison of primers were collected under protocol 20-0662, approved by the University of Colorado Boulder Institutional Review Board.

## Acknowledgements

We thank the University of Colorado Boulder for their ambitious and aggressive approach to keeping our campus safe. In particular, we thank Kristen Bjorkman for her key role in coordinating the pandemic response efforts on our campus. We thank Dan Larremore for valuable advice on the manuscript. This study is dedicated to the memory of our wonderful colleague, Denise Muhlrad.

